# Resonant breathing improves self-reported symptoms and wellbeing in people with Long COVID

**DOI:** 10.1101/2024.03.25.24304856

**Authors:** Jessica Polizzi, Jenna Tosto-Mancuso, Laura Tabacof, Jamie Wood, David Putrino

## Abstract

**Introduction:** Long COVID involves debilitating symptoms, many of which mirror those observed with dysautonomia, and care must be taken with traditional autonomic rehabilitation to avoid post-exertional malaise/post-exertional symptom exacerbation. Resonant breathing exercises require less exertion and can potentially improve autonomic function. The objective of this work was to report on the impact of a resonant breathing program on self-reported symptoms and wellbeing in people with Long COVID.

**Methods:** A retrospective analysis of de-identified data was completed in a convenience sample of people with Long Covid, who participated in the Meo Health (formerly known as Stasis HP) resonant breathing program. Participants completed baseline and follow up surveys.

**Results:** Data were available for 99 participants. Most measures of symptoms and wellbeing improved at follow up, with the largest differences per participant seen in sense of wellness (47.3%, p<0.0001), ability to focus (57.5%, p<0.0001), ability to breathe (47.5%, p<0.0001), ability to control stress (61.8%, p<0.0001) and sleep quality (34.9%, p=0.0002). Most (92%) participants reported improvement at follow up on the Patient Global Impression of Change Scale.

**Conclusion:** Self-reported symptoms and wellbeing improved in people with Long COVID completing resonant breathing. Resonant breathing can be considered as an option within the broader treatment plan of people with Long COVID.

## Introduction

Post-acute sequelae of SARS-CoV-2 infection (PASC), also known as Long COVID, is a post-acute infection syndrome that involves debilitating symptoms and affects at least 6% (almost 20 million) of the adult US population.^1^ These persistent symptoms include post-exertional malaise, fatigue, weakness, pain, shortness of breath, cognitive dysfunction, sleep disturbances, fevers, and gastrointestinal issues, and affect every organ system.^2-4^ Further, many symptoms of Long COVID mirror those observed with dysautonomia, including orthostatic intolerance, palpitations, tachycardia, syncope, labile blood pressure, dizziness, and exercise intolerance.^5^

Orthostatic intolerance and postural orthostatic tachycardia syndrome (POTS) can commonly follow acute viral infections, and are similarly observed in up to 80% of people with Long COVID.^5-8^ Prior to the COVID-19 pandemic, approximately 50% of individuals with POTS reported a history of viral infection prior to symptom onset, suggesting this as a potential trigger.^9^ The presence of autonomic dysfunction in Long COVID is possibly due to mechanisms such as viral- or immune-mediated disruption of the autonomic nervous system,^10^ brainstem signaling abnormalities,^11^ and/or small fiber neuropathy.^12^ In addition, sympathetic mediated hypocapnia has been reported in people with autonomic dysfunction,^13^ and in Long COVID in the absence of hyperventilation,^14^ with an overlap observed in the symptoms of hypocapnia and Long COVID.

While autonomic rehabilitation can be of benefit to some people with Long COVID, care must be taken to avoid post-exertional malaise/post-exertional symptom exacerbation (PEM/PESE).^6^ Encouraging people with Long COVID to engage in physical activity that exceeds their exertional tolerance has been a commonly identified cause of worsening symptoms in self-report surveys.^2-3^ Furthermore, there is evidence to suggest that overexertion of people with Long COVID who have PEM/PESE can lead to local and global physiological damage.^15^ As such, strategies that may regulate autonomic nervous system activity with minimal exertion should be investigated for feasibility and efficacy in the Long COVID community. Resonant breathing exercises require less exertion than traditional autonomic rehabilitation approaches, and have previously been demonstrated to alleviate hypocapnia and balance sympathetic and parasympathetic activation.^16^ The physiological benefit of resonant breathing is underpinned by increased responsiveness of the baroreflex,^17-18^ leading to improved homeostatic control of blood pressure and heart rate, alongside increased parasympathetic activation.^16^ This could theoretically serve to counterbalance abnormally increased sympathetic nervous system activity that is known to occur in Long COVID.^19^

Consensus guidelines for the treatment of autonomic dysfunction in Long COVID encourage clinicians to identify and address the most disabling symptoms.^6,20^ Given the potential ability of resonant breathing exercises to improve the function of the autonomic nervous system, including this as part of the broader approach to autonomic rehabilitation has merit. The primary objective of this work was to report on the impact of an online resonant breathing program on self-reported symptoms and wellbeing in people with Long COVID.

## Methods

### Study Design

This was a retrospective analysis of de-identified data from a convenience sample of people with Long Covid, who participated in the Meo Health (formerly known as Stasis HP) resonant breathing program between September 2020 and November 2021. This study was determined to be exempt from the need for ethics review by the Mount Sinai Program for the Protection of Human Subjects due to the de-identified nature of the data (STUDY-24-00246).

### Participants

People aged 18 years or older with Long COVID were either referred to the Meo Health program via their care provider, or self-referred to the program.

### Resonance breathing protocol

Participants underwent an initial 4-week progressive resonant breathing program using a 4:6 (inspiratory:expiratory) second cadence. Participants were instructed via recorded video content, and advised to complete the resonant breathing exercises twice per day for five days per week (upon waking and in the evening prior to sleep). Instructions for nasal unblocking were also provided. The time of the sessions were initially set at 10 minutes in length, increasing up to 30 minutes by the end of week four as tolerated. Participants were advised to avoid caffeine or other stimulants prior to the sessions. All participants had the option to request an online supervised session to check technique, but this was not mandatory. Following the initial 4 weeks, participants were encouraged to continue the program for 12 weeks and beyond, as a mainstay part of their broader Long COVID treatment plan.

### Outcome survey

A survey designed for the program by Meo Health was administered prior to commencement and at a follow up date according to the length of time the participant used the program. The survey collected basic demographic data (gender and age), and self-reported symptoms and wellbeing on a Likert scale (1 to 5, 1 = Very low, 5 = Very high), including stress, ability to control stress, anxiety, sleep quality, breathlessness, ability to breathe, fatigue, ability to focus, and sense of wellness. The first survey also included self-reported results from the max exhale test (MET) (amount of time it takes the participant to exhale as slowly as possible after full nasal inspiration) and breath hold test (BHT) (time able to hold breath after normal inhalation), while the follow up survey included the Patient Global Impression of Change (PGIC), and how many times per week and minutes per session the resonant breathing was performed.

### Statistical analysis

Statistical analyses were undertaken with Stata (StataCorp, Stata Statistical Software Release: V.14). Data were analyzed using descriptive statistics, t-tests and pairwise correlation. Categorical data are presented as frequencies and proportions. Continuous data are presented as mean and standard deviation (SD), median and interquartile range (IQR) or median and range. Bonferroni’s adjustment was applied post t-tests (n=9) to provide a corrected alpha of 0.0056. Level of significance was set to 0.05 for all other tests.

## Results

Data were available for 99 participants (78 [78%] female, aged median [IQR] 49 [41 to 57] years) who were enrolled in the program and completed both baseline and follow up surveys. The median time at follow up was 95 (29 to 378) days. Baseline MET was 28 (16 to 47) seconds, and BHT was 41 (28 to 50) seconds. Participants reported completing the resonant breathing program 7 (5 to 8) times per week, for 8 (8 to 13) minutes per session.

Nearly all (89%) measures of symptoms and wellbeing improved at follow up, with the largest mean (SD) percentage improvement per participant observed in ability to control stress (61.8 [96.0]%, p<0.0001), ability to focus (57.5 [76.3]%, p<0.0001), ability to breathe (47.5 [82.3]%, p<0.0001), sense of wellness (47.3 [67.5]%, p<0.0001), and sleep quality (34.9 [72.5]%, p=0.0002) (Table 1). Most (92%) participants reported some level of improvement at follow up on the Patient Global Impression of Change Scale (Figure 1). Improvement in ability to control stress was positively correlated with the number of times the Meo Health program was used each week (r=0.38; p<0.001). Improvement in ability to control stress was also associated with reduced self-reported anxiety (r=-0.29; p=0.003) and improved sleep (r=0.30; p=0.003).

**Table 1.** Self-reported symptoms and wellbeing for all participants (n=99).

|  | Baseline | Follow up | Mean diff.<br>(95% CI) | Change per<br>participant (%) | P value* |
| --- | --- | --- | --- | --- | --- |
| Stress | 3.25 (1.01) | 2.95 (0.97) | -0.29 (-0.57 to -0.02) | -2.7 (37.9) | 0.0389 |
| Ability to control stress | 2.39 (1.09) | 3.16 (0.97) | 0.77 (0.50 to 1.06) | 61.8 (96.0) | <0.0001 |
| Anxiety | 3.62 (1.11) | 2.66 (0.98) | -0.61 (-0.90 to -0.32) | -9.5 (48.0) | 0.0001 |
| Sleep quality | 2.81 (1.02) | 3.35 (0.97) | 0.55 (0.27 to 0.82) | 34.9 (72.5) | 0.0002 |
| Breathlessness | 2.67 (1.03) | 2.09 (1.05) | -0.57 (-0.87 to -0.28) | -14.1 (45.8) | 0.0001 |
| Ability to breathe | 2.81 (0.95) | 3.61 (0.90) | 0.80 (0.54 to 1.06) | 47.5 (82.3) | <0.0001 |
| Fatigue | 3.85 (1.12) | 3.28 (1.04) | -0.57 (-0.87 to 0.26) | -3.8 (62.8) | 0.0003 |
| Ability to focus | 2.53 (0.94) | 3.45 (0.82) | 0.93 (0.68 to 1.18) | 57.5 (76.3) | <0.0001 |
| Sense of wellness | 2.14 (0.86) | 2.79 (0.80) | 0.65 (0.41 to 0.88) | 47.3 (67.5) | <0.0001 |

Data are presented as mean (SD) unless otherwise stated.
\*Bonferroni's adjustment was applied post t-tests (n=9) to provide a corrected alpha of 0.0056.

**Figure 1.**
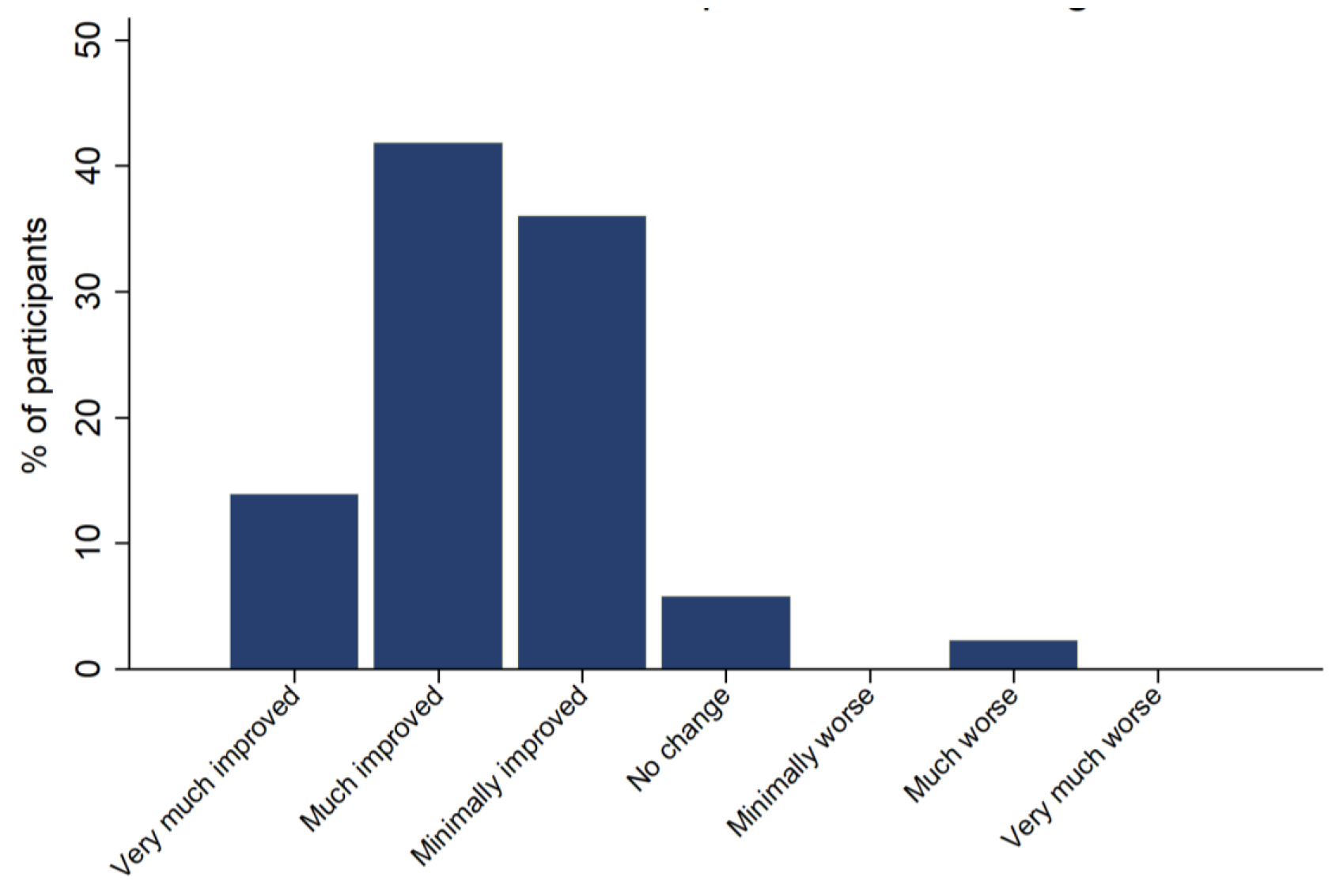
Patient Global Impression of Change scores for all participants (n=99).

## Discussion

The emergence of Long COVID has presented significant challenges to healthcare and the scientific community, with no approved treatments currently available (pharmacological or non-pharmacological). These data provide insight into the potential role of resonant breathing in the alleviation of some of the symptoms of Long COVID, alongside an ability to improve in self-reported measures of wellbeing in a small cohort.

The positive outcomes observed in people with Long COVID performing at least 4 weeks of resonant breathing support the hypothesis of improved autonomic function, aligning with previous findings highlighting the potential role of breathwork as a therapy to address autonomic dysfunction in Long COVID^21^ and other conditions.^22-24^ Controlled breathing may improve autonomic function via improving the baroreflex,^17-18^ enhancing parasympathetic activity and mitigating stress responses.^25^ Symptoms of autonomic dysfunction feature heavily in many presentations of Long COVID,^2-3,10^ therefore the reported data are encouraging. Resonant breathing is a relatively simple technique that can be easily integrated into the treatment programs of people with Long COVID.

In addition, using heart rate variability (HRV) biofeedback may further optimize the benefits of resonant breathing,^26^ warranting further investigation. HRV biofeedback can be integrated into smartphone applications and is therefore accessible for use by people in the home setting. The feasibility of using HRV biofeedback has been demonstrated in a small number (13) of people with Long COVID, with observed improvements in measures of symptoms and wellbeing.^21^

There are several limitations to be considered with this report. The data were analyzed retrospectively from a small cohort of people with Long COVID, with no prospective study design or control group. Information regarding how long each participant had been experiencing symptoms of Long COVID was not obtained, making it difficult to determine any level of benefit relating to how soon resonant breathing was commenced after initial infection. Further, while it is presumed that the participants were completing the program consistently up until the time at follow up, the data are self-reported and there may be discrepancies between responses and actual rates of adherence to the program. Other therapies being used were also not considered and therefore could not be controlled for in the analysis. The results now need to be reproduced in a larger prospective study with adequate controls.

This study provides some insight into the potential role and benefit of resonant breathing, and its feasibility as a simple and easy to use non-pharmacological treatment option for Long COVID. Long COVID impacts multiple body systems and requires a multi-targeted treatment approach,^27^ and a low-exertion therapy for improving autonomic nervous system function has the potential to be of benefit for many people with Long COVID.

## Data Availability

All data produced in the present study may be made available upon reasonable request to the authors

## References

1. Adjaye-Gbewonyo D, Vahratian A, Perrine CG, et al. Long COVID in adults: United States, 2022. NCHS Data Brief, no 480. Hyattsville, MD: National Center for Health Statistics. 2023. DOI: 10.15620/cdc:132417.

2. Davis HE, Assaf GS, McCorkell L, et al. Characterizing long COVID in an international cohort: 7 months of symptoms and their impact. EClinicalMedicine. 2021 Aug;38:101019. doi: 10.1016/j.eclinm.2021.101019. Epub 2021 Jul 15. PMID: 34308300; PMCID: PMC8280690.

3. Tabacof L, Tosto-Mancuso J, Wood J, et al. Post-acute COVID-19 Syndrome Negatively Impacts Physical Function, Cognitive Function, Health-Related Quality of Life, and Participation. Am J Phys Med Rehabil. 2022;101(1):48–52. doi:10.1097/PHM.0000000000001910

4. Klein J, Wood J, Jaycox JR, et al. Distinguishing features of long COVID identified through immune profiling. Nature. 2023;623(7985):139–148. doi:10.1038/s41586-023-06651-y

5. Larsen NW, Stiles LE, Miglis MG. Preparing for the long-haul: Autonomic complications of COVID-19. Auton Neurosci. 2021;235:102841. doi:10.1016/j.autneu.2021.102841

6. Blitshteyn S, Whiteson JH, Abramoff B, et al. Multi-disciplinary collaborative consensus guidance statement on the assessment and treatment of autonomic dysfunction in patients with post-acute sequelae of SARS-CoV-2 infection (PASC). PM&R. 2022; 14(10): 1270–1291. doi:10.1002/pmrj.12894

7. Eldokla AM, Mohamed-Hussein AA, Fouad AM, et al. Prevalence and patterns of symptoms of dysautonomia in patients with long-COVID syndrome: A cross-sectional study. Ann Clin Transl Neurol. 2022 Jun;9(6):778–785. doi: 10.1002/acn3.51557. Epub 2022 Apr 8. PMID: 35393771; PMCID: PMC9110879.

8. Haloot J, Bhavaraju-Sanka R, Pillarisetti J, et al. Autonomic Dysfunction Related to Postacute SARS-CoV-2 Syndrome. Phys Med Rehabil Clin N Am. 2023;34(3):563–572. doi:10.1016/j.pmr.2023.04.003

9. Sandroni P, Opfer-Gehrking TL, McPhee BR, et al. Postural tachycardia syndrome: clinical features and follow-up study. Mayo Clin Proc. 1999;74(11):1106–1110. doi:10.4065/74.11.1106

10. Dani M, Dirksen A, Taraborrelli P, et al. Autonomic dysfunction in ‘long COVID’: rationale, physiology and management strategies. Clin Med (Lond). 2021 Jan;21(1):e63–e67. doi: 10.7861/clinmed.2020-0896. Epub 2020 Nov 26. PMID: 33243837; PMCID: PMC7850225.

11. Proal AD, VanElzakker MB. Long COVID or Post-acute Sequelae of COVID-19 (PASC): An Overview of Biological Factors That May Contribute to Persistent Symptoms. Front Microbiol. 2021;12:698169. Published 2021 Jun 23. doi:10.3389/fmicb.2021.698169

12. Novak P, Mukerji SS, Alabsi HS, et al. Multisystem Involvement in Post-Acute Sequelae of Coronavirus Disease 19. Ann Neurol. 2022;91(3):367–379. doi:10.1002/ana.26286

13. Stewart JM, Pianosi P, Shaban MA, et al. Postural Hyperventilation as a Cause of Postural Tachycardia Syndrome: Increased Systemic Vascular Resistance and Decreased Cardiac Output When Upright in All Postural Tachycardia Syndrome Variants. J Am Heart Assoc. 2018;7(13):e008854. Published 2018 Jun 30. doi:10.1161/JAHA.118.008854

14. Wood J, Tabacof L, Tosto-Mancuso J,et al. Levels of end-tidal carbon dioxide are low despite normal respiratory rate in individuals with long COVID. J Breath Res. 2021;16(1):10.1088/1752-7163/ac3c18. Published 2021 Dec 8. doi:10.1088/1752-7163/ac3c18

15. Appelman, B., Charlton, B.T., Goulding, R.P. et al. Muscle abnormalities worsen after post-exertional malaise in long COVID. Nat Commun 15, 17 (2024). 10.1038/s41467-023-44432-3

16. Jerath R, Edry JW, Barnes VA, et al. Physiology of long pranayamic breathing: neural respiratory elements may provide a mechanism that explains how slow deep breathing shifts the autonomic nervous system. Med Hypotheses. 2006;67(3):566–71. doi: 10.1016/j.mehy.2006.02.042. Epub 2006 Apr 18. PMID: 16624497.

17. Sakakibara M, Kaneda M, Oikawa LO. Efficacy of Paced Breathing at the Low-frequency Peak on Heart Rate Variability and Baroreflex Sensitivity. Appl Psychophysiol Biofeedback. 2020 Mar;45(1):31–37. doi: 10.1007/s10484-019-09453-z. PMID: 31781925.

18. Lin G, Xiang Q, Fu X, et al. Heart rate variability biofeedback decreases blood pressure in prehypertensive subjects by improving autonomic function and baroreflex. J Altern Complement Med. 2012 Feb;18(2):143–52. doi: 10.1089/acm.2010.0607. PMID: 22339103.

19. Astin R, Banerjee A, Baker MR, et al. Long COVID: mechanisms, risk factors and recovery. Exp Physiol. 2023 Jan;108(1):12–27. doi: 10.1113/EP090802. Epub 2022 Nov 22. PMID: 36412084.

20. Herrera JE, Niehaus WN, Whiteson J, et al. Multidisciplinary collaborative consensus guidance statement on the assessment and treatment of fatigue in postacute sequelae of SARS-CoV-2 infection (PASC) patients [published correction appears in PM R. 2022 Jan;14(1):164]. PM R.2021;13(9):1027-1043. doi:10.1002/pmrj.12684

21. Corrado J, Iftekhar N, Halpin S, et al. HEART Rate Variability Biofeedback for LOng COVID Dysautonomia (HEARTLOC): Results of a Feasibility Study. Adv Rehabil Sci Pract. 2024 Jan 28;13:27536351241227261. doi: 10.1177/27536351241227261. PMID: 38298551; PMCID: PMC10826406.

22. Mourya M, Mahajan AS, Singh NP, et al. Effect of slow- and fast-breathing exercises on autonomic functions in patients with essential hypertension. J Altern Complement Med. 2009 Jul;15(7):711–7. doi: 10.1089/acm.2008.0609. PMID: 19534616.

23. Garg P, Mendiratta A, Banga A, et al. Effect of breathing exercises on blood pressure and heart rate: A systematic review and meta-analysis. Int J Cardiol Cardiovasc Risk Prev. 2023 Dec 27;20:200232. doi: 10.1016/j.ijcrp.2023.200232. PMID: 38179185; PMCID: PMC10765252.

24. Herawati I, Mat Ludin AF M M, et al. Breathing exercise for hypertensive patients: A scoping review. Front Physiol. 2023 Jan 25;14:1048338. doi: 10.3389/fphys.2023.1048338. PMID: 36760529; PMCID: PMC9905130.

25. Russo MA, Santarelli DM, O’Rourke D. The physiological effects of slow breathing in the healthy human. Breathe (Sheff). 2017 Dec;13(4):298–309. doi: 10.1183/20734735.009817. PMID: 29209423; PMCID: PMC5709795.

26. Lehrer PM, Gevirtz R. Heart rate variability biofeedback: how and why does it work? Front Psychol. 2014 Jul 21;5:756. doi: 10.3389/fpsyg.2014.00756. PMID: 25101026; PMCID: PMC4104929.

27. Nalbandian, A., Sehgal, K., Gupta, A. et al. Post-acute COVID-19 syndrome. Nat Med 27, 601–615 (2021). 10.1038/s41591-021-01283-z

